# Rehabilitation clinicians’ perspectives of reactive balance training

**DOI:** 10.1101/2021.02.19.21252082

**Authors:** David Jagroop, Stephanie Houvardas, Cynthia J Danells, Jennifer Kochanowski, Esmé French, Nancy M. Salbach, Kristin Musselman, Elizabeth L. Inness, Avril Mansfield

## Abstract

**Purpose:** Reactive balance training (RBT) aims to improve reactive balance control. However, because RBT involves clients losing balance, clinicians may view that it is unsafe or not feasible for some clients. We aimed to explore how clinicians are specifically implementing RBT to treat balance and mobility issues.

**Materials and methods:** Physiotherapists and kinesiologists across Canada who reported that they include RBT in their practices were invited to complete telephone interviews about their experience with RBT. Interviews were transcribed verbatim, and analysed using a deductive thematic analysis.

**Results:** Ten participants completed telephone interviews, which lasted between 30-60 minutes. Participants were primarily working in a hospital setting (inpatient rehabilitation (n=3); outpatient rehabilitation (n=2), and were treating those with neurological disorders (n=5). Four main themes were identified: 1) there is variability in RBT approaches; 2) knowledge can be a barrier and facilitator to RBT; 3) reactive balance control is viewed as an advanced skill; and 4) RBT experience builds confidence.

**Conclusions:** Our findings suggest a need for resources to make clinical implementation of RBT more feasible.

## INTRODUCTION

In Canada, falls are the most common cause of injury that require hospitalizations [1]. A fall occurs when there is a loss of balance, where individuals lose control of the relationship between their centre of mass and base of support. The ability to prevent a fall following a loss of balance depends on effective reactive balance control.

Reactive balance training (RBT) is the only type of exercise that has been shown to significantly improve reactive balance control [2-7]. RBT is a novel type of exercise that allows individuals to experience an intentional loss of balance, with the use of repeated balance perturbations in a safe environment. Those that complete RBT demonstrate improved control of stepping and reach-to-grasp reactions [2,7-13], and report fewer falls in daily life [14].

Clinicians are uncertain whether RBT is feasible in practice, due to the perception that RBT requires specialized moving platforms [2] or expensive programmable treadmills [15]. Our previous study reported that 75% of Canadian physiotherapists and kinesiologists who treat people for balance problems use RBT in their practices [16]. However, from that survey study it was unclear how clinicians were specifically implementing RBT. Additionally, some responses to the questionnaire suggested that clinicians equated volitional step training with RBT. There were also mixed opinions regarding the equipment needed for RBT. Thus, qualitative methods are warranted to gain further in-depth understanding of clinician’s use of RBT in practice. Therefore, the primary purpose of the current study was to explore how clinicians are specifically implementing RBT to treat balance and mobility issues. The secondary purpose was to gain a more in-depth understanding of the underlying barriers and facilitators to implementing RBT in practice.

## METHODS

The current study employed a qualitative descriptive approach using interviews to gain an in depth understanding of clinician’s use of RBT in practice. The standards for reporting qualitative research were followed [17]. Physiotherapists and kinesiologists that treat clients in any setting for problems with balance and mobility were invited to participate in the study. Respondents to our previous nation-wide survey study [16] who expressed an interest in participating in future research and who reported that they conducted RBT in their practices were invited to participate. Results from this study also informed our sampling approach; that is, we aimed to ensure our study sample was representative of the population of Canadian clinicians who treat people with balance problems in terms of, gender identity (80% women; 20% men), profession (80% physiotherapist; 20% kinesiologist), and number of years of experience (at least n=2 from each category). After providing informed consent, participants were asked to complete a short screening questionnaire about their profession, practice setting, client population, and use of RBT in practice.

One-on-one telephone interviews were conducted using a semi-structured interview guide, which was developed based on findings from our previous work [16] (**table 1**). For familiarization purposes, participants were sent a list of the interview questions approximately one week before the scheduled interview. Participants were invited to share additional thoughts, if any, after the interview via e-mail. Interviews were conducted by author DJ, who has previous experience in RBT research, and conducting interviews and focus groups.

**Table 1:**
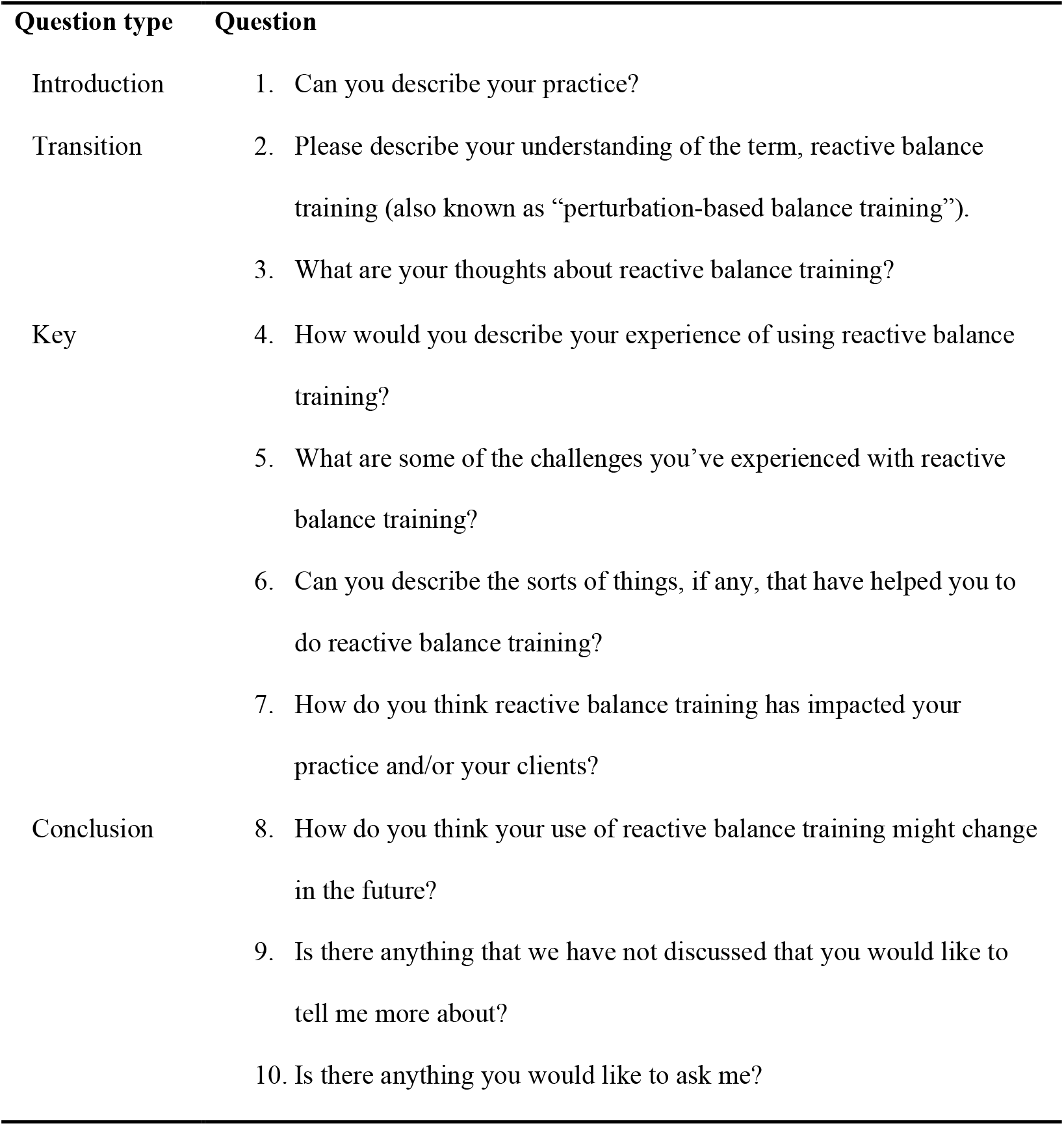
Interview guide.

| Question type | Question |
| --- | --- |
| Introduction | 1. Can you describe your practice? |
| Transition | 2. Please describe your understanding of the term, reactive balance training (also known as “perturbation-based balance training”). |
|  | 3. What are your thoughts about reactive balance training? |
| Key | 4. How would you describe your experience of using reactive balance training? |
|  | 5. What are some of the challenges you’ve experienced with reactive balance training? |
|  | 6. Can you describe the sorts of things, if any, that have helped you to do reactive balance training? |
|  | 7. How do you think reactive balance training has impacted your practice and/or your clients? |
| Conclusion | 8. How do you think your use of reactive balance training might change in the future? |
|  | 9. Is there anything that we have not discussed that you would like to tell me more about? |
|  | 10. Is there anything you would like to ask me? |

Telephone interviews were audio recorded and later transcribed verbatim. Field notes provided additional context to the responses, and thoughts that were shared post-interview (via e-mail) were added to the interview transcripts. A deductive thematic analysis of the transcripts was performed using a codebook, which included codes that had emerged from previous work [16]. As our previous study used a questionnaire to learn about clinical practices around RBT, a deductive thematic analysis was employed to gain a more in-depth understanding of current practices [18]. One transcript was selected and coded individually by 3 coders (AM, DJ, and SH). The coders selected quotes that supported codes. The 3 coders then met as a group to discuss their findings, as well as any discrepancies with coding (i.e., which code was most appropriate for a particular quote). All coders reached a mutual consensus on the revised codebook, and the remaining transcripts were analyzed by the 3 coders. The final codebook was then revised and condensed into major theme summaries. Finally, an executive summary of the key findings was created.

## RESULTS

Characteristics of the ten participants are presented in **table 2**. Interviews typically lasted between 30 to 60 minutes. One interview had technical difficulties with recording audio, and as a result, the latter half of the interview was not recorded. Field notes by the interviewer were used in place of the missing portion of the interview. Two participants provided additional comments via e-mail, which were added to their transcripts. A total of 4 main themes were identified from the interviews: 1) there is variability in RBT approaches; 2) knowledge can be both a barrier and facilitator to RBT; 3) reactive balance control is viewed as an advanced skill; and 4) RBT experience builds confidence. Verbatim quotes were used to support study findings. Themes, and sub-themes from the interviews are presented in **table 3**.

**Table 2:**
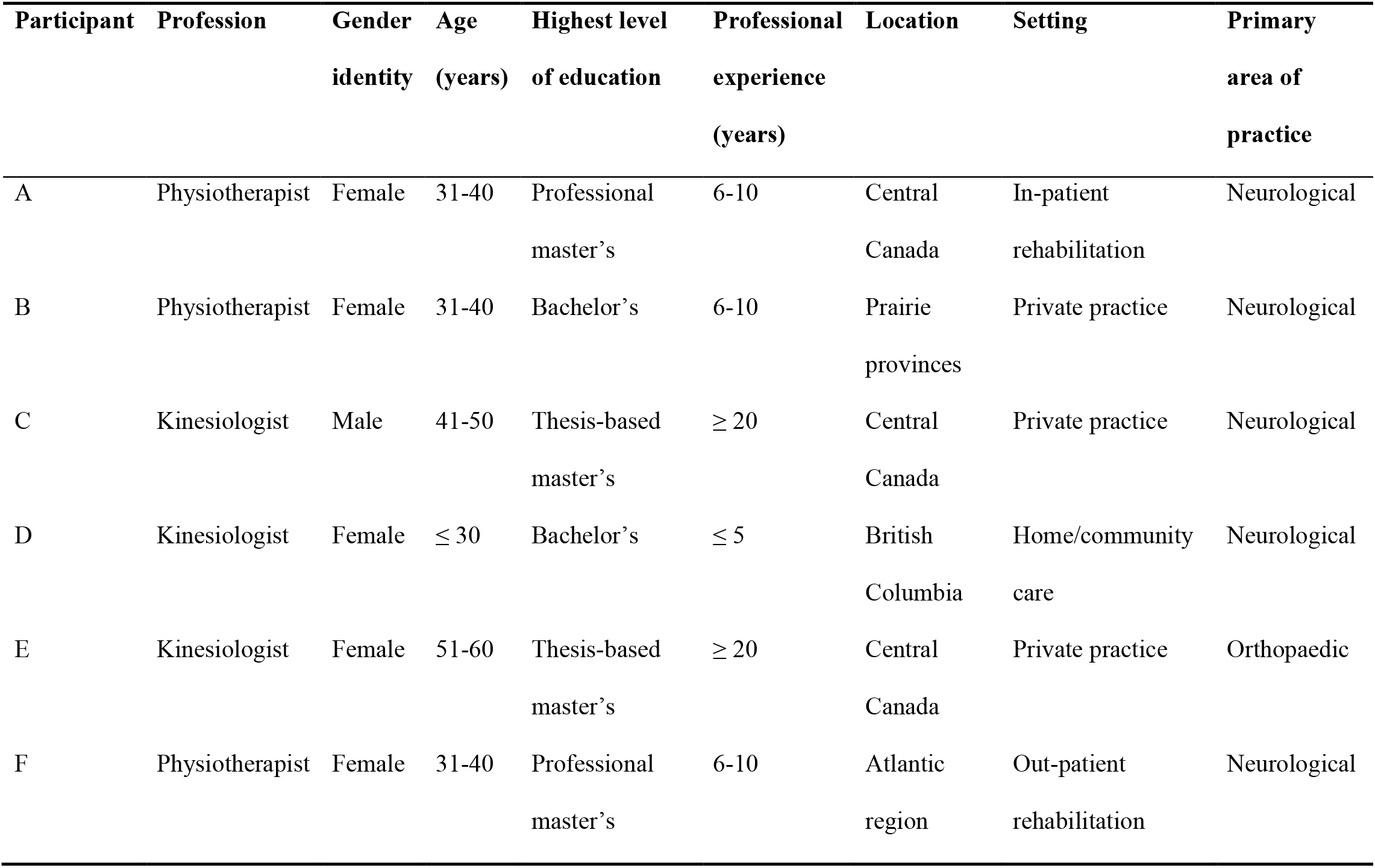
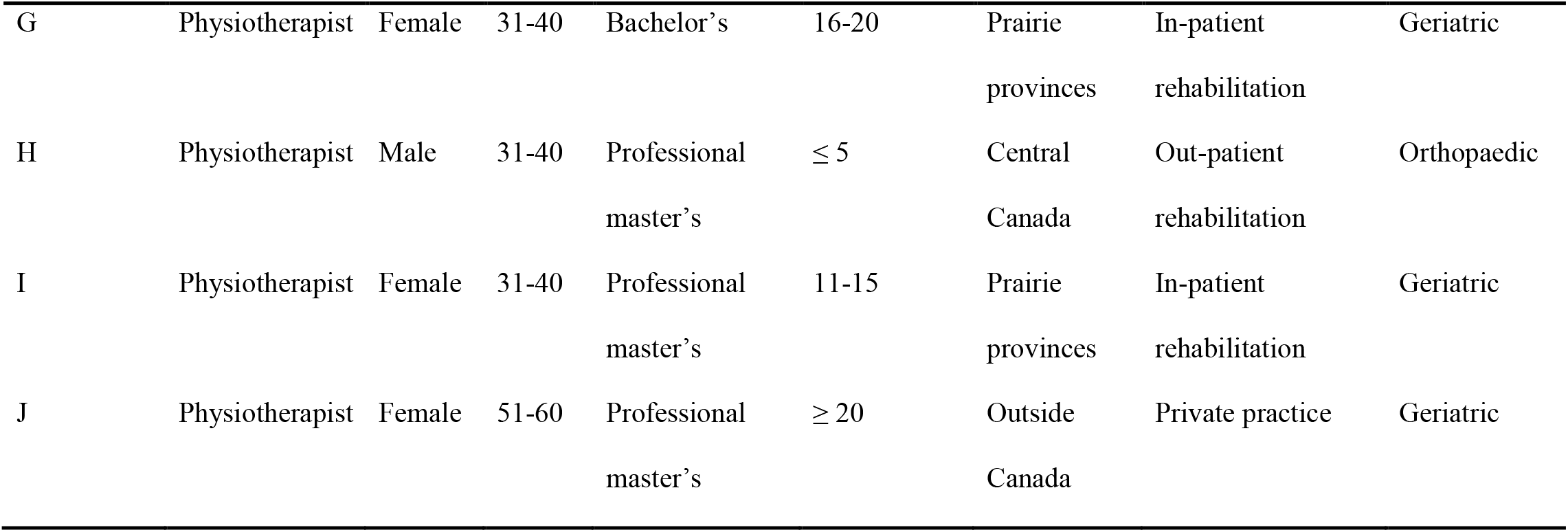
Participant characteristics. Values presented as number of responses in each category.

| Participant | Profession | Gender<br>identity | Age<br>(years) | Highest level<br>of education | Professional<br>experience<br>(years) | Location | Setting | Primary<br>area of<br>practice |
| --- | --- | --- | --- | --- | --- | --- | --- | --- |
| A | Physiotherapist | Female | 31-40 | Professional<br>master's | 6-10 | Central<br>Canada | In-patient<br>rehabilitation | Neurological |
| B | Physiotherapist | Female | 31-40 | Bachelor's | 6-10 | Prairie<br>provinces | Private practice | Neurological |
| C | Kinesiologist | Male | 41-50 | Thesis-based<br>master's | ≥ 20 | Central<br>Canada | Private practice | Neurological |
| D | Kinesiologist | Female | ≤ 30 | Bachelor's | ≤ 5 | British<br>Columbia | Home/community<br>care | Neurological |
| E | Kinesiologist | Female | 51-60 | Thesis-based<br>master's | ≥ 20 | Central<br>Canada | Private practice | Orthopaedic |
| F | Physiotherapist | Female | 31-40 | Professional<br>master's | 6-10 | Atlantic<br>region | Out-patient<br>rehabilitation | Neurological |
| G | Physiotherapist | Female | 31-40 | Bachelor's | 16-20 | Prairie<br>provinces | In-patient<br>rehabilitation | Geriatric |
| H | Physiotherapist | Male | 31-40 | Professional<br>master's | $\leq 5$ | Central<br>Canada | Out-patient<br>rehabilitation | Orthopaedic |
| I | Physiotherapist | Female | 31-40 | Professional<br>master's | 11-15 | Prairie<br>provinces | In-patient<br>rehabilitation | Geriatric |
| J | Physiotherapist | Female | 51-60 | Professional<br>master's | $\geq 20$ | Outside<br>Canada | Private practice | Geriatric |

**Table 3:** Common themes, categories and sub-categories from interviews.

| Themes | Categories | Sub-categories |
| --- | --- | --- |
| 1. There is variability in RBT approaches | • Participants' definition of RBT | • High-level balance training vs evoking balance reactions |
|  | • Assessment approaches | • Limitations of current assessment strategies |
|  | • Treatment approaches | • RBT should be done face-to-face and one-on-one<br>• Lack of sufficient space<br>• Low-tech equipment used<br>• "You can get creative"<br>• Enjoyment for both the client and therapist<br>• Desire to have high-tech equipment<br>• Conflicts with treatment planning |
| 2. Knowledge can be a barrier and facilitator to RBT | • Lack of RBT knowledge/skills | • Uncertainty with applying RBT to practice<br>○ Appropriate clientele<br>○ When to introduce it into care<br>○ RBT prescription<br>○ Best methods to conduct RBT |
|  | • Prior knowledge of neuroscience facilitated RBT practice | • Greater understanding principles of RBT and the importance |
|  | • RBT research viewed as barrier to implementation | • Equipment used in research studies vs what is feasible<br>• Populations in research studies vs clients treated |
|  | • Learning needs for RBT practice | • How to assess reactive balance control<br>• How to implement RBT<br>• Creative ways to incorporate it without high-tech equipment |
| 3. Reactive balance control is viewed as an advanced skill | • RBT not appropriate for lower-functional clients (low motor or cognitive function) | • Priority of restoring function (ADLs)<br>• RBT at the end of treatment program<br>• ‘Running out of time’<br>• RBT as preventative therapy |
|  | • Applying RBT at home is not appropriate |  |
|  | • RBT as preventative therapy | • Need for RBT earlier in therapy |
|  | • Safety harnesses | • Concern of safety and lower-functioning clients<br>• Limitations to using harness |
|  | Human resources | Use of assistants in place of safety harness<br>Alleviating safety concerns<br>Access to an assistant and role of an assistant |
| 4. RBT experience builds confidence | Apprehensive/fearful about RBT | External perturbations |
|  | Increased self-confidence with practice |  |
|  | Trust between therapist and client |  |
|  | Client's feelings of RBT | Apprehensive/fearful/anxious<br>Benefits and enjoyment<br>Self-efficacy/sense of achievement |
|  | Team support | Facilitator<br>Barrier due to attitudes of RBT |

### There is variability in RBT approaches

#### Participants’ definition of RBT

Most participants defined RBT similar to “*the use of internally or externally executed perturbations to challenge a variety of reactions that simulates daily balance scenarios or specific balance deficits” (Participant I)*. However, some considered that RBT involves volitional step training, reacting to other stimuli (e.g., visual), or training to “maintain” balance when experiencing external perturbations rather than practicing reactions to recover balance after the perturbation.

> *“I’m standing around them… and do little perturbations, like a little push, they don’t know what direction its coming from*… *it’s from the side, the front, the back…. And they have to <u>maintain</u> their balance” (Participant B; emphasis added)*.

#### Assessment approaches

Although most participants understood that RBT involves training reactive balance control, participants did not use a standardized reactive balance assessment. The most commonly used assessment tools were the Berg Balance Scale and the Timed Up and Go. A few participants highlighted that existing clinical balance assessment tools were limited in their ability to assess reactive balance control.

> *“…you could have two clients that both get 56 out of 56 on their Berg balance, but one might be way better with reactive balance than the other*.*” (Participant B)*.

As they were not aware of a formal tool to assess reactive balance control, a few participants improvised and created their own methods or used movement observation as a way to assess balance problems.

> *“I could put the client in a situation like you know just standing on a carpet, or standing with their eyes closed. So changing, you know, assessing the same movement in the context of different stimuli or different conditions*.*” (Participant H)*.

#### Treatment approaches

Most participants used low-tech methods to cause clients to lose balance; for example, they used manual (external forces) or internal perturbations (client loses balance while completing a challenging balance task), with/without equipment that is typically found in a rehabilitation setting. Participants also mentioned incorporating games to make RBT more enjoyable. Some participants expressed interest in acquiring specialized equipment to provide perturbations (e.g., commercially available moving platforms), but also reported that lack of space and funding were barriers to acquiring such equipment. Others felt they had adequate equipment to provide RBT. Some participants made their own equipment such as moveable platforms, or used commonly found equipment (e.g., resistance bands) to evoke balance reactions. One participant’s institution had dedicated space for RBT, but difficulties scheduling access to the space posed challenges in conducting RBT. Participants expressed two major conflicts with incorporating RBT into their treatment plan: 1) discharge planning within their setting, and 2) client goals. With discharge planning, participants felt that their setting prioritized functional outcomes, which hindered their ability to do RBT. Indeed, some participants also mentioned that balance, in general, is not prioritized in therapy; rather, the main focus is independence with activities of daily living, and walking with an aid.

> *“I’m still working on explaining to our management team that balance is a priority to be part of that safe and functional discharge” (Participant I)*.

Many participants felt that clients also do not prioritize balance training.

> *“Certainly we have clients who sometimes think like “this is not what we are here for, this is a waste of my time, get me walking and then I’m good” …The balance part tends to fall away in favor of the, you know, is their walking speed at a safe community level or distance and things like that” (Participant I)*.

### Knowledge can be both a barrier and facilitator to RBT

Few participants expressed that they did not have experience and training in providing RBT. Most participants did not receive formal training with RBT in their professional education. Consequently, participants did not always know how and when to use RBT optimally. Participants were particularly unsure about how to prescribe RBT, when to conduct RBT in therapy, which clients were appropriate for RBT, and what RBT methods would be best for their clients.

> *“What’s the right amount and in what stage in their recovery can we use it? Should we just be doing internal? Should we just be doing external? Should we do a combination? Should we be doing strength, aerobics with it or just static, dynamic? I think that’s the part as a clinician I don’t have that answer… I think the challenging part is, how do you make it fit for the client, their abilities and actually challenge them enough that it makes a difference and they’re going to improve?” (Participant F)*.

Participants wanted better ways to assess reactive balance control, and more education on how to prescribe RBT. Some participants noted that their colleagues lack awareness of RBT.

> *“We need more education on it, or more creative ways to use it…. And I think the reason why it isn’t (implemented more in rehabilitation) is because not a lot of people know about it*.*” (Participant D)*.

In contrast, three participants felt that they had a strong background in neuroscience, motor control, and/or motor learning. These participants reported that this knowledge gained from their graduate and undergraduate education helped them better understand the principles of RBT and the importance of improving reactive balance control.

> *“Knowledge of motor control, motor learning, how skills are acquired, that kind of knowledge has helped me immensely understand reactive balance training. I did go back to school and did a Master’s degree where I did study balance training and motor skill acquisition and that helped me immensely understanding what I had been doing for the last 6 years. So more education certainly helped me understand it and helped me understand why I was doing some of the things I was doing at the clinic*.*” (Participant C)*.

While most participants noted that lack of knowledge was a barrier to RBT, and better education and training around RBT could improve implementation, one participant who was familiar with RBT research reported this as a barrier to implementation. This participant felt that the equipment-based RBT approaches used in research were not feasible in clinical practice, and that the participants included in RBT studies did not reflect the complexity of her clientele.

> *“When I read the studies on it, I believe researchers are not targeting anyone who looks like my patients, and that’s why I’m always in this dilemma… when the researcher shares in that study the demographics of the patients, my patients don’t look like those patients. Another barrier, you cannot do exactly what you guys are doing in a lab, in a clinic, because we don’t even have close to the same equipment*… *it’s not the same*.*” (Participant J)*.

### Reactive balance control is viewed as an advanced skill

Participants often viewed reactive balance control as an advanced skill. Consequently, many participants felt that RBT is not appropriate for lower-functioning clients (i.e., those with low motor function and/or cognitive function). Participants prioritized re-training function such as sitting or quiet standing balance with these types of clients, and tended to conduct RBT towards the end of clients’ treatment programs. This often meant that they ‘ran out of time’ to do RBT as clients were often discharged before they were ready for more ‘advanced’ balance training.

> *“I would like to say in a perfect world I would do a perturbed balance assessment on everyone before I create their balance program. But the reality is that most of my people would not be appropriate for that really formal kind of balance training or balance assessments when they first arrive because they simply can’t even stand up from a chair without assistance*.*” (Participant I)*.

In contrast, some participants mentioned that RBT should be done earlier in therapy rather than waiting until function is improved, and should also be used as a preventative therapy for those who do not yet have major balance problems/falls.

Participants frequently expressed that, since reactive balance control is an advanced skill, it is not appropriate for home exercise. Likewise, many participants felt RBT needs to be face-to-face and one-on-one; the need to constantly closely interact with the client during RBT was seen as a barrier.

> “*…it ties you up with that client, you just can’t give them something and walk away, so it has to be one-on-one…you have to be with the client there” (Participant E)*.

#### Safety harnesses

Participants expressed concern conducting RBT safely, particularly with lower-functioning clients. Participants that had access to a safety harness stated that they used the harness to prevent the client from falling to the floor. However, some participants noted that they would sometimes only use this system in the beginning of RBT, and then later progress to training without it.

> *“I don’t do harnesses unless I’m doing that really firm push and if it’s usually in their early stages where they’re a little bit more fearful and I don’t know if they’re going to lose their balance, so I like to be safe. After I know they can do it we get rid of the harnesses*.*” (Participant F)*.

A few participants felt that the safety harness either changed the client’s balance reactions, or felt that clients became too reliant on it and would be afraid to train without it.

> *“I really rarely use that with my clients ‘cause I feel like it actually changes the way they are receiving the input. So they ended up relying on the harness and not actually relying on how, um, how their own proprioception or weight-bearing should be providing feedback to the brain*.*” (Participant I)*.

#### Human resources

Most participants did not have access to a safety harness system, and as an alternative, either did RBT in parallel bars or had another individual (e.g., another clinician or a therapy assistant) assist to spot the client. The need for another person to assist with RBT presented a barrier; participants reported difficulty with scheduling assistants during training times. Furthermore, as reactive balance control was viewed as an advanced skill, and therefore that they viewed RBT as complex, participants did not feel comfortable delegating RBT entirely to a therapy assistant.

### RBT experience builds confidence in both the client and therapist

Some participants expressed that they were apprehensive and fearful about doing RBT, specifically external perturbations; however, practice increased their self-confidence and self-efficacy to do it with more clients.

> *“So I think with each person you feel like a little more confident. Maybe a little bit more like hey, I do have a good sense of things and I can be a little more creative on how I challenge this person or make what we’re doing in this really artificial setting relevant to their everyday life, which would help” (Participant A)*

Likewise, participants reported that clients often expressed apprehension, fear, and anxiety, but they also acknowledged the benefits of RBT and enjoyed the training. Some participants noted that RBT improves clients’ self-efficacy because they experience a sense of achievement from doing something challenging.

> *“Reactive balance training, if it’s done well, can give the client that’s doing it a feeling of achievement and a feeling of success that they attempted something that was difficult and had some success in it, and that can drive sort of further success and further progress in rehabilitation” (Participant C)*.

Because of clients’ fear and anxiety with doing RBT, the trust between clinician and client was also noted to be important.

> *“I think building rapport which is something I felt that I’ve done really well with my clients in explaining the purpose of it… so they understand like the importance and how it’s going to get them to where they want to be… um, then, just having that support makes the engagement with them a lot better” (Participant D)*.

## DISCUSSION

With a goal of increasing clinical uptake of this important therapy, the current study explored how clinicians use RBT to treat balance issues, and their perspectives on implementing RBT in practice. We found that there is variability in definitions and approaches to RBT, knowledge can be both a barrier and facilitator to RBT, reactive balance control is viewed as an advanced skill, and experience with RBT can improve confidence for therapists and clients.

Reactive balance training involves participant repeatedly experiencing loss of so that they can practice, and improve control of, balance reactions. However, findings from our previous study [16] and the current study suggest that some participants consider RBT to be volitional step training, reacting to other stimuli (e.g., visual stimuli), or practicing maintaining rather than recovering balance after external perturbations. This finding highlights the need for researchers to clearly report the ‘active ingredients’ of their interventions, using checklists such as the Template for Intervention Description and Replication [20]. Likewise, clinicians are responsible for delivering evidence-based interventions that, as closely as possible, replicate the intervention that was studied [21]. Researchers should develop clinician-friendly training guidelines to aid in this clinical implementation. Furthermore, while there is no evidence that activities such as volitional step training can improve reactive balance control [22], reactive and volitional step training have not been directly compared [23]. Since volitional step training is used by clinicians, and is likely more feasible to implement than RBT, there is likely value in studying the effect of volitional step training on reactive balance control.

Interestingly, one participant viewed knowledge of RBT research as a barrier to clinical implementation. The two predominant reasons being that they perceived: 1) RBT research subjects were less medically complex and higher functioning than those seen in clinical practice; and 2) RBT research uses high-tech equipment that is not available for clinical use. Likewise, many participants felt that reactive balance control is an advanced skill, and therefore RBT is not appropriate for lower-functioning clients. Previous work has found that, while clinicians recognize the need for challenging balance exercises, clinicians are unlikely to prescribe such challenging exercises, especially in group settings, due to concerns of clients falling [24]. As a result, clients tend not to exercise at or near the limits of postural stability [25]. This finding highlights the need to include study participants with a range of medical and functional characteristics and, when diverse research participants are included, to more clearly report the diversity of these characteristics (e.g., by reporting ranges for baseline assessments, in addition to means and standard deviations). This finding also highlights the need for clinically-feasible training approaches to be used in clinical trials of RBT. Consistent with our previous survey study, participants disagreed as to whether specialized equipment is required for RBT [16].

Participants mentioned that being creative and improvising with equipment that was already available in clinic enabled RBT, without need for high-tech equipment. Several research studies have demonstrated that RBT can be conducted using internal and/or manual perturbations with the use of low-tech or no equipment [7-9,11,26-28].

Participants reported limitations of existing standardized tools for assessing reactive balance control, and consequently used informal observation of clients’ balance to inform treatment. Assessments such as the Berg Balance Scale and Timed Up and Go were often used by participants in our study, and are commonly used in other settings [29-31]. However, these tools do not include items related to reactive balance control, and it seems that participants were unaware of tools such as the Balance Evaluation Systems Test (BEST) [32] or the Performance Oriented Mobility Assessment (POMA) [33], which do include reactive balance control items. This finding is consistent with other studies showing that reactive balance control is assessed infrequently in clinical practice [31], and that rehabilitation clinicians do not often use standardized tools with reactive balance control components, such as the BEST or POMA, to assess balance control [34]. When clinicians do assess reactive balance control, it is usually by movement observation rather than using a standardized tool [35].

Participants’ uncertainties with using RBT in practice fostered apprehensive feelings, particularly with the use of external perturbations. Conversely, participants noted that experience increased confidence in RBT. Therefore, if therapists consistently include RBT in routine practice, this can build confidence and self-efficacy [36]. Participants also mentioned their clients’ feelings of apprehension, fear, and anxiety, but also feelings of enjoyment and improved self-efficacy as the training progressed. Although clients’ feelings about RBT were reported indirectly, these reports are consistent with Pak and colleagues [37]; which, also found that client’s were able to overcome their fear and anxiety about reactive balance assessments, due to their trust in their therapist. Pak et al. [37] and our current study emphasize the importance of building trust with clients, creating a positive rapport, and applying a graded approach in order to find an appropriate challenge for each client [38]. Having the client involved in the training process, and listening to their preferences can help promote feelings of success and ultimately enhance the overall RBT experience for both the client and therapist [38,39].

## Data Availability

Not applicable.

## LIMITATIONS

This study investigated the perspectives of rehabilitation clinicians that use a very specific type of balance training (RBT). Although purposeful sampling was used to seek those that conduct RBT, these individuals may have a greater interest in this training, and may be more inclined to participate to share their positive experiences. Participants also mentioned their clients’ perspective of RBT; these perspectives might not actually represent client perspectives.

## CONCLUSION

Overall, these findings can support implementation strategies to increase the use of RBT in practice, which in turn, can reduce the risk of falls across multiple populations. Specifically, the study findings suggest a need for: 1) guidelines that clearly define the ‘active ingredients’ of RBT; 2) educational tools to guide use of RBT in practice (e.g., when to use RBT and with which clients); and 3) trust between the therapist and client when conducting RBT.

